## Supplemental information 1 for "Incidence of Lyme disease in the United Kingdom and association with fatigue: a population-based, historical cohort study"

Supplementary information 1 - List of comorbidities

| Comorbidity/condition | Source of code lists | | Number of participants excluded with this comorbidity | |
| --- | --- | --- | --- | --- |
| Comorbidities associated with long-term fatigue (= any record prior index as exclusion criteria) | | | | |
| Lupus | | IQVIA | | 8 |
| Polymyositis | | IQVIA | | 0 |
| Heart failure | | IQVIA | | 16 |
| Idiopathic pulmonary fibrosis | | IQVIA | | 1 |
| Severe COPD | | IQVIA | | 5 |
| Chronic kidney disease | | (1) | | 39 |
| Diabetes | | (1) | | 61 |
| Addison’s or Cushing’s diseases | | IQVIA | | 1 |
| Multiple sclerosis | | IQVIA | | 7 |
| Parkinson’s disease | | IQVIA | | 3 |
| Narcolepsy | | IQVIA | | 0 |
| Rheumatoid arthritis | | IQVIA | | 15 |
| Comorbidities/conditions associated with short-term fatigue (= any record within 12 months prior index as exclusion criteria) | | | | |
| Hypothyroidism/ Hyperthyroidism | | Adapted from (2) | | 16 |
| Cancer | | IQVIA | | 10 |
| Severe anaemia | | IQVIA | | 3 |
| Severe enduring mental illness | | (1) | | 1 |
| Anorexia | | Adapted from (2) | | 0 |
| Sleep apnoea | | IQVIA | | 3 |
| Hepatitis | | IQVIA | | 0 |
| Tuberculosis | | IQVIA | | 1 |
| Alcoholism | | IQVIA | | 8 |
| Pregnancy | | IQVIA | | 22 |

1.	Finnikin S, Ryan R, Marshall T. Cohort study investigating the relationship between cholesterol, cardiovascular risk score and the prescribing of statins in UK primary care: study protocol. BMJ Open. 2016 Nov 1;6(11):e013120.

2.	University of Cambridge. CPRD @ Cambridge – Codes Lists [Internet]. 2018. Available from: https://www.phpc.cam.ac.uk/pcu/research/research-groups/crmh/cprd_cam/codelists/v11/
