## Supplemental information 2 for "Incidence of Lyme disease in the United Kingdom and association with fatigue: a population-based, historical cohort study"

Supplementary information 2 - Covariates

Age at index was categorized in 10-year wide categories, except for children under 15 and adults above 75. BMI was directly extracted or, when missing, calculated from weight and height. Value on the latest date before or on index date was extracted, considering only records within 1 year before index for patients under 18. BMI was grouped into 4 categories according to UK National Health Service classification (underweight, healthy weight, overweight, and obese). Smoking status was directly extracted from the data as 3 categories (never smoker, ex-smoker, current smoker) or, when missing, supplemented using specific read codes (1). Healthcare Utilisation Frequency was calculated as the number of face-to-face and phone consultations within 6 months before index and categorized into 5 categories: 0, 1, 2-4, 5-9, and more than 10. Index season was created based on index date as 4 categories: Jan/Feb/March, April/May/June, July/August/Sept, and Oct/Nov/Dec. Antibiotic treatment was a binary variable based on the presence of any record of Amoxicillin, Azithromycin, [Cefotaxime](https://www.drugs.com/mtm/cefotaxime.html), Ceftriaxone or Doxycycline, i.e. antibiotics recommended in the UK for the treatment of Lyme disease (2), +/-31 days of index date, or on or within 31 days of confirmed diagnosis date. The rationale behind the inclusion of the pre-diagnosis period stems from NICE recommendations to start treatment while waiting for test results (2). History of depression was a binary variable based on the presence of any record of mild and moderate depression-related codes.

1.	Finnikin S, Ryan R, Marshall T. Cohort study investigating the relationship between cholesterol, cardiovascular risk score and the prescribing of statins in UK primary care: study protocol. BMJ Open. 2016 Nov 1;6(11):e013120.

2.	NICE. Lyme disease NICE guideline [Internet]. 2018. Available from: https://www.nice.org.uk/guidance/ng95
