## Supplemental information 3 for "Incidence of Lyme disease in the United Kingdom and association with fatigue: a population-based, historical cohort study"

Supplementary information 3 - Incidence rates and hazard ratios (HR) of any types of fatigue and of chronic fatigue syndrome for study covariates (N=10,640)

|  | Any types of fatigue^a^ | | | | | Chronic fatigue syndrome | | | | |
| --- | --- | --- | --- | --- | --- | --- | --- | --- | --- | --- |
| Baseline Characteristics | Number of events | Total person-years | Incidence rate^b^ | HR^c^ (95% CI) | p value^h^ | Number of events | Total person-years | Incidence rate^b^ | HR^d^ (95% CI) | p value^h^ |
| **Overall** | 929 | 48,228 | 192.62 |  |  | 17 | 51,700 | 3.29 |  |  |
| **Sex** |  |  |  |  |  |  |  |  |  |  |
| Female | 573 | 22,951 | 249.66 | Not assessed^e^ |  | 8 | 25,127 | 3.18 | 0.93 (0.36-2.41) | 0.880 |
| Male | 356 | 25,277 | 140.84 | Not assessed^e^ |  | 9 | 26,572 | 3.39 | ref |  |
| **Age (in years)** |  |  |  |  |  |  |  |  |  |  |
| <15 | 51 | 6266 | 81.4 | Not assessed^e^ |  | 0 | 6,441 | 0.00 | Not assessed^f^ |  |
| 15-24 | 56 | 2815 | 198.94 | Not assessed^e^ |  | 1 | 3,017 | 3.31 | 0.38 (0.05-3.07) | 0.366 |
| 25-34 | 67 | 4000 | 167.49 | Not assessed^e^ |  | 2 | 4,267 | 4.69 | 0.57 (0.12-2.70) | 0.480 |
| 34-44 | 183 | 8830 | 207.24 | Not assessed^e^ |  | 2 | 9,605 | 2.08 | 0.27 (0.06-1.26) | 0.095 |
| 45-54 | 201 | 9506 | 211.44 | Not assessed^e^ |  | 8 | 10,233 | 7.82 | ref |  |
| 55-64 | 211 | 10271 | 205.42 | Not assessed^e^ |  | 4 | 11,040 | 3.62 | 0.47 (0.14-1.57) | 0.220 |
| 65-74 | 115 | 5296 | 217.13 | Not assessed^e^ |  | 0 | 5,704 | 0.00 | Not assessed^f^ |  |
| >=75 | 45 | 1244 | 361.72 | Not assessed^e^ |  | 0 | 1,392 | 0.00 | Not assessed^f^ |  |
| **Body Mass Index**^g^ | |  |  |  |  |  |  |  |  |  |
| Underweight | 12 | 626 | 191.83 | 0.95 (0.44-2.04) | 0.892 | 1 | 674 | 14.84 | 3.44 (0.41-28.57) | 0.253 |
| Healthy weight | 301 | 14,538 | 207.05 | ref |  | 6 | 15,658 | 3.83 | ref |  |
| Overweight | 264 | 12,205 | 216.30 | 0.99 (0.80-1.21) | 0.886 | 6 | 13,170 | 4.56 | 1.18 (0.38-3.66) | 0.775 |
| Obese | 180 | 6,387 | 281.82 | 1.38 (1.09-1.74) | 0.007 | 2 | 7,044 | 2.84 | 0.70 (0.14-3.46) | 0.660 |
| **History of depression** | |  |  |  |  |  |  |  |  |  |
| No | 663 | 40,856 | 162.28 | ref |  | 10 | 43,463 | 2.30 | ref |  |
| Yes | 266 | 7,373 | 360.79 | 1.99 (1.66-2.39) | < 0.001 | 7 | 8,237 | 8.50 | 3.54 (1.34-9.29) | 0.010 |
| ^a^Symptoms of fatigue, post-viral fatigue, or chronic fatigue syndrome; ^b^per 10,000 person-years; ^c^using Cox regression with adjustment for the match variable; ^d^using Cox regression with no adjustment; ^e^already adjusted via the match variable; ^f^data sparsity; ^g^N=7,597; underweight<18.5; healthy weight:18.50-24.99; overweight 25-29.99; obese≥ 30.00, in kg/m^2^; ^h^Wald test for specific categories *continued* | | | | | | | | | | |

Incidence rates and rate ratios (HR) of any types of fatigue and of chronic fatigue syndrome for study co-variates (N=10,640) *(continued)*

|  | Any types of fatigue^a^ | | | | | Chronic fatigue syndrome | | | | |
| --- | --- | --- | --- | --- | --- | --- | --- | --- | --- | --- |
| Baseline Characteristics | Number of events | Total person-years | Incidence rate^b^ | HR^c^ (95% CI) | p value^g^ | Number of events | Total person-years | Incidence rate^b^ | HR^d^ (95% CI) | p value^g^ |
| **Antibiotic treatment**^e^ | | | | | | | | | | |
| No | 701 | 40,848 | 171.61 | ref |  | 11 | 43,430 | 2.53 | ref |  |
| Yes | 228 | 7,381 | 308.91 | 2.15 (1.81-2.56) | < 0.001 | 6 | 8,270 | 7.25 | 2.84 (1.05-7.68) | 0.040 |
| **Healthcare Utilisation Frequency**^f^ | | | | | | | | | | |
| None | 198 | 18,229 | 108.62 | ref |  | 4 | 19,001 | 2.11 | ref |  |
| 1 | 144 | 9,584 | 150.25 | 1.63 (1.27-2.09) | < 0.001 | 4 | 10,130 | 3.95 | 1.89 (0.47-7.57) | 0.366 |
| 2 to 4 | 306 | 13,032 | 234.81 | 2.52 (2.03-3.12) | < 0.001 | 4 | 14,255 | 2.81 | 1.29 (0.32-5.17) | 0.717 |
| 5 to 9 | 194 | 5,752 | 337.25 | 4.34 (3.37-5.59) | < 0.001 | 2 | 6,430 | 3.11 | 1.40 (0.26-7.65) | 0.698 |
| 10+ | 87 | 1,631 | 533.37 | 7.23 (5.15-10.15) | < 0.001 | 3 | 1,884 | 15.93 | 7.03 (1.56-31.65) | 0.011 |
| **Index season** | | | | | | | | | | |
| Jan. to March | 79 | 3,904 | 202.34 | Not assessed^h^ |  | 4 | 4,218 | 9.48 | Not assessed^h^ |  |
| April to June | 209 | 10,354 | 201.86 | Not assessed^h^ |  | 3 | 11,118 | 2.70 | Not assessed^h^ |  |
| July to Sept. | 441 | 23,161 | 190.40 | Not assessed^h^ |  | 9 | 24,802 | 3.63 | Not assessed^h^ |  |
| Oct. to Dec. | 200 | 10,810 | 185.02 | Not assessed^h^ |  | 1 | 11,562 | 0.86 | Not assessed^h^ |  |
| **Smoking status**^i^ | | | | | | | | | | |
| Never | 433 | 21,546 | 200.97 | ref |  | 7 | 23,192 | 3.02 | ref |  |
| Ex | 221 | 8,735 | 253.01 | 1.39 (1.14-1.70) | 0.001 | 6 | 9,490 | 6.32 | 2.05 (0.69-6.10) | 0.197 |
| Current | 197 | 9,429 | 208.94 | 1.32 (1.07-1.64) | 0.011 | 4 | 10,193 | 3.92 | 1.33 (0.39-4.56) | 0.649 |
| ^a^Symptoms of fatigue, post-viral fatigue, or chronic fatigue syndrome^; b^per 10,000 person-years; ^c^using Cox regression with adjustment for the match variable; ^d^using Cox regression with no adjustment; ^e^record of antibiotics used against Lyme disease within 30 days of index date or within 30 days after confirmed diagnosis for Lyme cohort patients, if different; ^f^number of GP encounters within 6 months prior index; ^g^Wald test for specific categories; ^h^given time since index was the time axis used for the Cox regression; ^i^N=8,887 | | | | | | | | | | |
